# Checklist to Support the Development and Implementation of AI in Clinical Settings

**DOI:** 10.1101/2024.08.08.24311701

**Authors:** Ayomide Owoyemi, Joanne Osuchukwu, Megan Salwei, Andy Boyd

**Affiliations:** University of Illinois Chicago, Chicago, IL, 60608, USA; University of Cincinnati College of Medicine, Cincinnati, OH, 45229, USA; Vanderbilt University Medical Center, Nashville, TN 37232, USA

**Keywords:** Artificial Intelligence, Sociotechnical, Checklist

## Abstract

The integration of Artificial Intelligence (AI) in healthcare settings demands a nuanced approach that considers both technical performance and sociotechnical factors. Recognizing this, our study introduces the Clinical AI Sociotechnical Framework (CASoF), developed through literature synthesis, and refined via a Modified Delphi study involving global healthcare professionals. Our research identifies a critical gap in existing frameworks, which largely focus on either technical specifications or trial outcomes, neglecting the comprehensive sociotechnical dynamics essential for successful AI deployment in clinical environments. CASoF addresses this gap by providing a structured checklist that guides the planning, design, development, and implementation stages of AI systems in healthcare. The checklist emphasizes the importance of considering the value proposition, data integrity, human-AI interaction, technical architecture, organizational culture, and ongoing support and monitoring, ensuring that AI tools are not only technologically sound but also practically viable and socially adaptable within clinical settings. Our findings suggest that the successful integration of AI in healthcare depends on a balanced focus on both technological advancements and the socio-technical environment of clinical settings. CASoF represents a step forward in bridging this divide, offering a holistic approach to AI deployment that is mindful of the complexities of healthcare systems. The checklist aims to facilitate the development of AI tools that are effective, userfriendly, and seamlessly integrated into clinical workflows, ultimately enhancing patient care and healthcare outcomes.

## 1 Introduction

The implementation of any technology in a real-world setting especially a clinical one requires adequate consideration of the social aspects of its application alongside the technical considerations.[1] The National Academy of Medicine report highlighted the need to “understand the technical, cognitive, social, and political factors in play and incentives impacting integration of AI into health care workflows”.[2] It is important to understand the context in which the technology will be used, how it will work with existing workflows without disruption and how it will be accepted by the people who will have to use it. Historically, in the development of Artificial Intelligence systems, the technical perspective has taken pre-eminence over how they fit and work in the real world, and this has resulted in AI systems falling short of their translational goals.[3] In general, AI tools have shown promise in development but few have been able to translate into the real world settings for patient management.[4] For example, a management decision tool built and deployed in a hospital in Utah for diabetes management had a challenge of not offering all the information that was desired by clinicians and patients to decide on Type 2 Diabetes (T2D) management.[5]

Despite the numerous proof-of-concept publications in this field, the lack of robust frameworks for supporting development and management of these tools is one of the main barriers to their adoption in healthcare.[6] There is a paucity of specific guidance and rigorous best practices for people designing and developing AI solutions targeted at clinical settings and use-cases. Findings from a review conducted by Gama et al highlighted the need to develop an AI-specific implementation framework because there is an unrealized opportunity to draw insights from implementation science, use theoretical and practical insights to accelerate and improve on the implementation of AI in clinical settings.[7]

There have been a few frameworks and guidelines proposed recently. Building on the work of Salwei et al., (2021), Salwei & Carayon developed a sociotechnical systems (STS) framework for AI that acknowledges the social and technical aspects of work that relate to the successful design and implementation of AI.[1] Their model demonstrates that an AI can only integrate in clinical workflows if it fits within the context, or the work system, in which it is implemented. Two other models, the Consolidated Standards of Reporting Trials-AI extension and Transparent reporting of a multivariable prediction model for individual prognosis or diagnosis (TRIPOD), are examples of models that are narrow in their application and focused on trials, performance and comparison which are only helpful in a single phase of the AI life cycle.[8], [9]

## 2 Methods

### 2.1 Literature Synthesis

In identifying sources for this literature synthesis, we searched multiple electronic databases which included MEDLINE via OVID, and Embase. This search was done between the 25^th^ and 30^th^ of June 2023. The literature search was focused on identifying articles that intersect the domains of artificial intelligence with the implementation of clinical frameworks, guidelines, theories, and their development, design, and evaluation. The following keywords were used in the search: “Artificial intelligence”, “Framework”, “Guideline”, “Theory”, “Implementation”, “Evaluation, “Design”, “Development”, “Clinical Settings”, “Clinical Care”, “Hospital”, “Clinic”, “Patient Care”. There were no date limitations. The search resulted in the identification of 573 potential studies. The abstracts of the retrieved studies were reviewed using the following inclusion and exclusion criteria:

a.) studies that involved the application of AI by healthcare providers in a clinical setting, b.) research that employed a conceptual or theoretical framework related to AI in clinical care and c.) primary qualitative studies concerning the design, implementation, or predictive evaluation of AI in clinical care, irrespective of the use of a distinct framework. While the exclusion criteria were a.) studies that primarily focused on patient-related outcomes, b.) research that concentrated solely on the technical performance or computational aspects of AI without clinical integration.

This process yielded 19 relevant studies which were then reviewed by reading the full articles. This resulted in the exclusion of three studies, one for being a reporting guideline, another for being a study protocol, and the third for being a commentary piece. I also examined the included articles and their citations and identified four other relevant studies to make a final total of 20 articles. These articles were reviewed, and the key points, themes and insights extracted. These were combined and synthesized with the results from a previously conducted primary study evaluating the implementation and user experience of an AI-powered sepsis alert system at a clinical facility.

### 2.2 The Modified Delphi Study

A preliminary checklist designed to assist teams in creating AI systems for clinical settings was developed from the literature synthesis. This checklist underwent a review, editing, and improvement process by a purposively selected group of experts using the Delphi method, a technique aimed at achieving consensus through systematic feedback from a panel of knowledgeable individuals. [10] This method is particularly useful in healthcare for establishing guidelines or protocols when evidence is scarce or conflicting. [11] The review process took place over nearly two months consisting of two rounds, targeting a wide range of healthcare professionals and researchers with at least two years of experience in their fields. The study had a global reach, and to avoid any panelist bias, the identities of the participants were kept anonymous, utilizing Google Forms for data collection.

The Delphi study involved an initial survey with questions rated on relevance to the AI system’s development process on a Likert scale (ranked 1 – 5), along with space for open-ended feedback. The checklist was organized into four key stages of AI system development, aligning with the framework’s six domains. 80 individuals showed interest, with 65 invited to participate, and 38 completing the first round. Feedback from this round led to revisions of the checklist for a subsequent round, focusing on questions deemed relevant by achieving a rating of 4 or higher. Analysis included descriptive statistics and an assessment of consensus, with a mean score of 0.8 (agreed apriori) indicating agreement among panelists. The reliability of the responses was evaluated using the Cronbach alpha coefficient, and qualitative feedback was analyzed to refine the checklist further, using Python for the quantitative analysis.

## 3 Results

### 3.1 Literature Synthesis

The search conducted identified 23 studies that either proposed a framework, guideline or approach for the design, development, implementation, or evaluation of Artificial Intelligence for clinical use cases. Most of these addressed specific areas in the AI development cycle from design to maintenance and management, while some cut across every aspect of the cycle. The results of the literature search were synthesized with the primary research to arrive at the domains of the **CLINICAL AI SOCIOTECHNICAL FRAMEWORK** which is a sociotechnical framework to support the planning, design, development, and proposed implementation of AI systems to help better plan and predict the likely success of the AI system (Appendix).

**Fig. 1.**
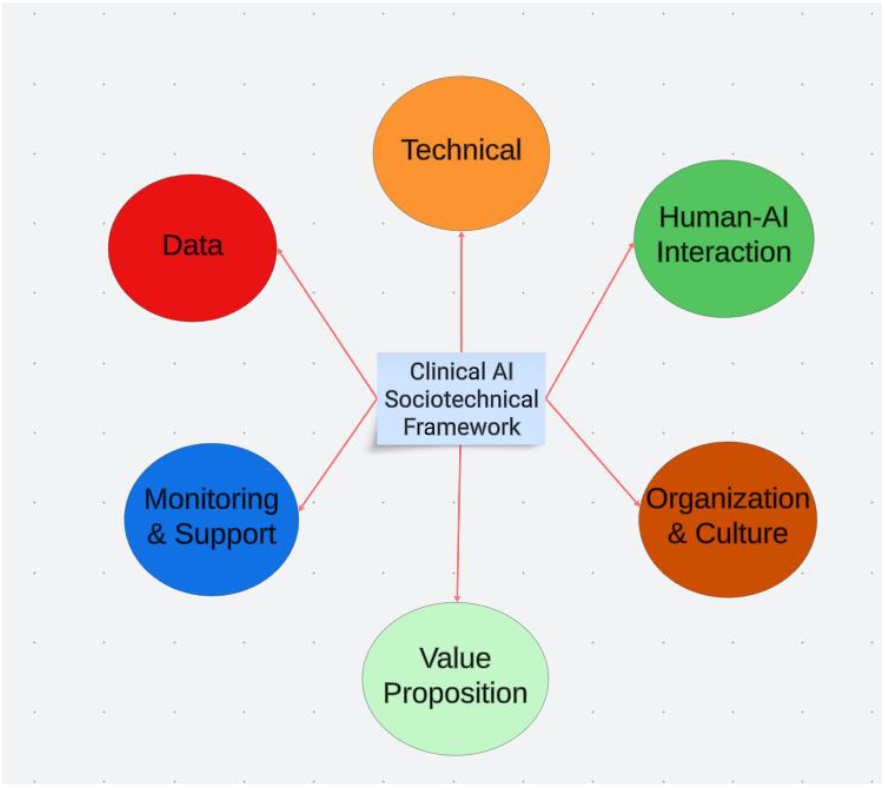
The Clinical AI Sociotechnical Framework

### 3.2 Literature Synthesis

Based on the Clinical AI Sociotechnical Framework, the first draft of the checklist was developed which was shared to a team of panelists for evaluation and review using a Delphi approach. A total of 64 panelists were recruited with 32.3% of them being Doctors, 15.4% were healthcare experts or researchers, AI researchers were 12.3%, health informaticians and Nurses were 6.2% respectively, and other professionals making up the rest. Of the 65 panelists invited to participate in the study, 38 of them completed the first round of the Delphi. The initial checklist had four overall categories which corresponded to the four stages in the development and deployment process, with 15 sub-categories which corresponded to the domains of the Clinical AI Sociotechnical Framework (CASoF) that were important in each of the stages. The stages were “Planning Stage”, “Design Stage”, “Development Stage” and “Proposed Implementation Stage”. As part of the questionnaire, panelists were asked two open ended questions at the end of each of the sub-categories which are “Would you reframe any of the questions above?” and “Are there questions that you would add or remove from this segment?”. During the first round of the Delphi, panelists suggested multiple edits and additions to the checklist. This suggested editing included the need to reframe some of the questions to make them more appropriate for a checklist and clearer. In one of the sub-categories, one panelist responded by saying “**The last question says ‘data processing’. That comes across as ambiguous. What does that refer to? who will be the audience for this survey? will they understand what that means? Are we trying to abstract curation, cleaning etc into abstraction?**”

At the end of the survey, panelists were asked why they might not use the checklist and some of the responses included: “**I think the checklist is long. The challenge when you have checklists this long is that people tend to gloss over them and are not intentional about answering the questions in a detailed way**”. Another in line with that comment also said “**Might be helpful to shorted and make more actionable. E.g policies and procedures document has been completed vs. have you considered a place for policies**” and another said “**The checklist is somewhat burdensome on the AI vendor and health system. I would cut the questions in half**”. These open-ended questions were analyzed using a content analysis approach to bring out the recurrent themes and perspectives shared by the panelists in reforming and improving the questionnaire. Quantitative analyses were done which showed a high level of agreement and relevance across most questions. Descriptive analysis was done. The mean score for relevance of the questions on the survey exceeded 0.8 on all but one, indicating that at least 80% of respondents found the questions pertinent to their work and the topic at hand. Furthermore, the interquartile range (IQR) was calculated to be 0 for all questions except three, highlighting a level of consensus among respondents. The consensus and the structure of the checklist is outlined below (Table 1).

**Table 1.** Summary of sub-categories by domain for round one.

| Stage | Domain | Number of Questions | Percentage of statements with Consensus |
| --- | --- | --- | --- |
| Planning Stage | Value Proposition | 4 | 75% (3) |
|  | Data | 4 | 100% (4) |
|  | People | 3 | 100% (3) |
|  | Organization and Culture | 3 | 100% (3) |
| Design Stage | User Interface and Experience | 2 | 100% (2) |
|  | Workflow | 4 | 100% (4) |
| Development Stage | Technical | 3 | 100% (3) |
|  | Clinical Utility | 4 | 75% (3) |
|  | Workflow | 2 | 50% (1) |
|  | Data | 2 | 100% (2) |
|  | User Interface and Experience | 3 | 100% (3) |
| **Proposed Implementation-Stage** | Organization and Culture | 1 | 100% (1) |
|  | People | 3 | 100% (3) |
|  | Technical | 3 | 100% (3) |
|  | Monitoring and Support | 4 | 100% (4) |

Note: Consensus was achieved when 80% of panelists rated a question as relevant Total number of participants: 38

Based on the results, comments, and feedback from the panelists the checklist was revised and edited. The Design and Development stages were merged into a single stage, the People and Organization and Culture domains were merged into a single domain. The user experience and workflow and clinical utility were merged to create a new domain called Human-AI Interaction. The total number of questions was reduced from 45 questions to 34 questions to make it less cumbersome and focused. These 34 questions were sent to the all the registered panelists for a second round of the Delphi. All the recruited panelists were included in the second and invited to review the updated checklist. Quantitative analyses were done which showed a high level of agreement and relevance across most questions. Descriptive analysis was done. The mean score for relevance of the questions was more than 0.8 on all questions, indicating that at least 80% of respondents found the questions pertinent to their work and the topic at hand. Furthermore, the interquartile range (IQR) was calculated to be 0 for all questions, highlighting a level of consensus among respondents. Based on the outcome of the Delphi a final checklist was outlined with one more question added to make 35 in total (Table 2).

**Table 2.** Final draft of the Clinical AI Sociotechnical Framework (CASoF) Checklist.

| Stage | Domain | Questions |
| --- | --- | --- |
| **Planning Stage** | Value Proposition & Utility | Have you outlined the expected impacts on patient outcomes?<br>Have you outlined its expected impact on care provider efficiency and outcomes?<br>Has any economic analysis been conducted for the AI system? |
|  | Data | Have you engaged in the use of any ethical data checklist during your data collection and preparation?<br>Have you engaged domain experts in the data preparation, cleaning, and engineering process?<br>Have you delineated an approach to maintain data quality, integrity, and security. |
|  | Human-AI Interaction | Have you accounted for existing clinical workflows and protocols? |
|  | Organization and Culture | Have you identified key stakeholders and their needs?<br>Have you identified potential resistance or barriers within the organization? |
|  |  | • Are there strategies in place to facilitate and ensure end-user engagement in the design and development phase?<br>• Do you have a good understanding of the culture within the institution, relevant department and teams and changes that might be needed? |
| **Design & Development Stage** | Technical | • Are you planning for hardware/software (EHR if relevant) systems and requirements?<br>• Are you creating support documentation for users and management e.g. model details, explainability details, data details, metrics, manuals etc.?<br>• Have you validated clinical accuracy and reliability?<br>• Have you secured any required regulatory approval?<br>• Have you taken active steps to mitigate against biased results? |
|  | Human-AI Interaction | • Have you conducted a simulation with end-users in real work system scenarios?<br>• Have you evaluated if the outputs are clear and understandable for the users?<br>• Have you implemented patient and user safety measures?<br>• Have you assessed the impact on the delivery of clinical tasks?<br>• Have you involved and tested with users?<br>• Has resistance to the use of the AI system been identified and addressed?<br>• Are you developing strategies to ensure that the alerts from the AI system are relevant, timely, and not overwhelming, to avoid alert fatigue? |
|  | Data | • Have you tested your method on various types of data to make sure it works well in different situations?<br>• Have you planned for data drift and shift (changes in the data over time)? |
| **Proposed Implementation Stage** | Organization and Culture | • Have you ensured that this intervention aligns with the existing governance and regulatory frameworks of the organization?<br>• Have you prepared necessary training/resources for end users?<br>• Have you considered steps to help address end users' questions and alleviate their concerns? |
|  | Technical | • Have you conducted a real-world evaluation of the AI system?<br>• Are you planning for pilot/silent tests?<br>• Are you providing user tools for continuous validation and evaluation of the system? |
|  | Monitoring and Support | • Have you created a plan to evaluate the success of the implementation? |
|  |  | • Have you planned for continuous user feedback on the system?<br>• Have you planned for regular audits, reviews, and updates?<br>• Have you planned for continuous education and support for users? |

## 4 Discussion

Enhancing the real-world impact of AI tools involves navigating a nuanced blend of technical and social elements. This process demands a strategic framework that guides the planning and preparation efforts throughout the AI tool’s life cycle, from its initial conceptualization to its sustained application. CASoF, based on its socio-technical perspective, encompasses different existing frameworks by providing a structured overview of the critical issues related to the integration, validation, and operationalization of AI in healthcare. CASoF offers a high-level approach to solving the translational and adoption problems bedevilling AI systems designed for clinical settings. CASoF can be used singly or in combination with some of the other existing frameworks in evaluating AI systems. Lennerz et al.’s Diagnostic Quality Model (DQM) and Jin et al.’s Clinical Explainable Artificial Intelligence (XAI) Guidelines, delve into diagnostic quality and explainability within medical imaging.[12], [13] They provide structured methodologies that could refine CASoF by integrating rigorous quality assessments and enhancing transparency in AI tools. The strengths of these frameworks lie in their focused criteria, which could synergistically enrich CASoF’s scope, ensuring that AI’s clinical implementation is both effective and socio-technically sound.

When comparing the Clinical AI Socio-technical Framework (CASoF) to the Stakeholder Engagement in the Development and Implementation of AI Technologies (SALIENT) framework and the Translational Evaluation of Healthcare AI (TEHAI) framework, distinct thematic focuses and approaches emerge, highlighting each framework’s unique contributions to the discourse on AI implementation in healthcare.[3], [14] CASoF stands out with its socio-technical checklist that integrates human, technical, and organizational culture in healthcare AI, while the SALIENT framework’s structured approach focuses more on the AI deployment lifecycle, including stakeholder engagement and ethical considerations, without delving as deeply into the sociotechnical interplay of CASoF. The TEHAI framework emphasizes the translational journey from AI development to clinical deployment, aligning with CASoF’s practical application but not matching its breadth in addressing socio-technical dynamics. These differences arise due to each framework’s underlying focus.

CASoF helps to create a practical plan shaped by the complex nature of healthcare in the real world as the successful implementation of AI depends on a mix of being ready at an organizational level, thinking ahead about the work environment, and involving all stakeholders. CASoF pushes for being adaptable and following a usercentered approach and ongoing interaction with stakeholders. This ensures AI tools are not just good technologically but also fit well socially and within organizations. It aims for AI to work smoothly with current health systems and to be sustainable in the long run, thinking about how AI can grow and be maintained from the start.

### 4.1 Limitations

While the primary research, literature synthesis and Delphi technique offer a robust approach to the development of the framework and checklist for development and integration of AI in the clinical setting, the real-world application would be more difficult and not as straightforward as the research might suggest. Therefore, there might be a need for continuous refinement of CASoF through iterative feedback and broader engagement with more stakeholders. Future research should aim to include an even wider array of perspectives, particularly from underrepresented regions and specialties, to enhance the framework’s comprehensiveness and applicability. The framework further encounters limitations in capturing the full spectrum of technical challenges, needs and their implications across diverse healthcare contexts globally. Considering these constraints, application of the framework will benefit from synergistic application with other existing frameworks.

## 5 Conclusion

The CASoF offers an approach to bridging the gap between the technical aspects of AI and how they can be best planned to fit and work in the clinical setting with a view to improving the impact it makes on clinical work and patient outcomes. It offers a structured strategy to mitigate challenges and obstacles in the development and implementation process. CASoF offers an advancement over previous frameworks and approaches by holistically encapsulating the technical and sociotechnical dimensions necessary for AI to thrive within the clinical space.

## Data Availability

All data produced in the present study are available upon reasonable request to the authors

## References

[1] M. E. Salwei and P. Carayon, “A Sociotechnical Systems Framework for the Application of Artificial Intelligence in Health Care Delivery,” Journal of Cognitive Engineering and Decision Making, p. 15553434221097357, May 2022, doi: 10.1177/15553434221097357.

[2] M. E. Matheny, D. Whicher, and S. Thadaney Israni, “Artificial Intelligence in Health Care: A Report From the National Academy of Medicine,” JAMA, vol. 323, no. 6, pp. 509–510, Feb. 2020, doi: 10.1001/jama.2019.21579.

[3] S. Reddy et al., “Evaluation framework to guide implementation of AI systems into healthcare settings,” BMJ Health Care Inform, vol. 28, no. 1, p. e100444, Oct. 2021, doi: 10.1136/bmjhci-2021-100444.

[4] J. He, S. L. Baxter, J. Xu, J. Xu, X. Zhou, and K. Zhang, “The practical implementation of artificial intelligence technologies in medicine,” Nat Med, vol. 25, no. 1, pp. 30–36, Jan. 2019, doi: 10.1038/s41591-018-0307-0.

[5] S. Tarumi et al., “Leveraging Artificial Intelligence to Improve Chronic Disease Care: Methods and Application to Pharmacotherapy Decision Support for Type-2 Diabetes Mellitus,” Methods Inf Med, vol. 60, no. S 01, pp. e32–e43, Jun. 2021, doi: 10.1055/s-0041-1728757.

[6] D. Ben-Israel et al., “The impact of machine learning on patient care: A systematic review,” Artificial Intelligence in Medicine, vol. 103, p. 101785, Mar. 2020, doi: 10.1016/j.artmed.2019.101785.

[7] F. Gama, D. Tyskbo, J. Nygren, J. Barlow, J. Reed, and P. Svedberg, “Implementation Frameworks for Artificial Intelligence Translation Into Health Care Practice: Scoping Review,” Journal of Medical Internet Research, vol. 24, no. 1, p. e32215, Jan. 2022, doi: 10.2196/32215.

[8] X. Liu, S. C. Rivera, D. Moher, M. J. Calvert, and A. K. Denniston, “Reporting Guidelines for Clinical Trial Reports for Interventions Involving Artificial Intelligence,” Lancet Digit Health, vol. 2, no. 10, pp. e537–e548, Oct. 2020, doi: 10.1016/S2589-7500(20)30218-1.

[9] G. S. Collins, J. B. Reitsma, D. G. Altman, and K. G. M. Moons, “Transparent reporting of a multivariable prediction model for individual prognosis or diagnosis (TRIPOD): the TRIPOD statement,” BMJ, vol. 350, p. g7594, Jan. 2015, doi: 10.1136/bmj.g7594.

[10] E. Taylor, “We Agree, Don’t We? The Delphi Method for Health Environments Research,” HERD, vol. 13, no. 1, pp. 11–23, Jan. 2020, doi: 10.1177/1937586719887709.

[11] E. Taylor, A. Joseph, X. Quan, and U. Nanda, “Designing a Tool to Support Patient Safety: Using Research to Inform a Proactive Approach to Healthcare Facility Design,” in Advances in Ergonomics In Design, Usability & Special Populations: Part III, AHFE Open Acces, 2022. doi: 10.54941/ahfe1001343.

[12] J. K. Lennerz et al., “Diagnostic quality model (DQM): an integrated framework for the assessment of diagnostic quality when using AI/ML,” Clinical Chemistry and Laboratory Medicine (CCLM), vol. 61, no. 4, pp. 544–557, Mar. 2023, doi: 10.1515/cclm-2022-1151.

[13] W. Jin, X. Li, M. Fatehi, and G. Hamarneh, “Guidelines and evaluation of clinical explainable AI in medical image analysis,” Medical Image Analysis, vol. 84, p. 102684, Feb. 2023, doi: 10.1016/j.media.2022.102684.

[14] A. H. van der Vegt, I. A. Scott, K. Dermawan, R. J. Schnetler, V. R. Kalke, and P. J. Lane, “Deployment of machine learning algorithms to predict sepsis: systematic review and application of the SALIENT clinical AI implementation framework,” J Am Med Inform Assoc, vol. 30, no. 7, pp. 1349–1361, May 2023, doi: 10.1093/jamia/ocad075.

